# Isolation of infected people and their contacts is likely to be effective against many short-term epidemics

**DOI:** 10.1101/2020.10.07.20207845

**Authors:** Nathan Geffen, Marcus Low

## Abstract

**Background:** Isolation of infected people and their contacts may be an effective way to control outbreaks of infectious disease, such as influenza and SARS-CoV-2. Models can provide insights into the efficacy of contact tracing, coupled with isolating or quarantining at risk people.

**Methods:** We developed an agent-based model and simulated 15, 000 short term illnesses, with varying characteristics. For each illness we ran ten simulations on the following scenarios: (1) No tracing or isolation (None), (2) isolation of agents who have tested positive (Isolation), (3) scenario 2 coupled with minimal contact tracing and quarantine of contacts (Minimum), (4) scenario 3 with more effective contact tracing (Moderate), and (5) perfect isolation of agents who test positive and perfect tracing and quarantine of all their contacts (Maximum).

**Results:** The median total infections of the Isolation, Minimum, Moderate and Maximum scenarios were 80%, 40%, 17% and 4% of the no intervention scenario respectively.

**Conclusions:** Isolation of infected patients and quarantine of their contacts, even if moderately well implemented, is likely to substantially reduce the number of infections in an outbreak. Randomized controlled trials to confirm these results in the real world and to analyse the cost effectiveness of contact tracing and isolation during coronavirus and influenza outbreaks are warranted.

## 1 Introduction

Several non-pharmaceutical interventions have been introduced in different countries to reduce the spread of the SARS-CoV-2 pandemic. Some of these have included lockdowns, physical distancing rules, prolific use of hand sanitisers, and mask-wearing [1, 2, 3].

In some countries, such as South Korea, a key intervention has been tracing the contacts of infected people so that they can isolate or quarantine themselves [4, 5]. Indeed, preparation for tracking coronavirus outbreaks was in place in South Korea long before the emergence of Covid-19 [6].

Isolation refers to the movement of people with the infection being restricted, voluntarily or legally, while quarantine refers to the movement of people at risk of infection being restricted, voluntarily or legally. The work described here does not differentiate between these distinctions. We therefore use *isolation* to refer interchangeably to isolation or quarantine, whether voluntarily undertaken or legally enforced.

Besides coronaviruses, contact tracing has also been implemented to control tuberculosis, hepatitis and HIV and other sexually transmitted infections [7, 8, 9, 10]. But these are rather different illnesses to Covid-19, generally infecting patients for longer and spreading slower through populations. It is also not usually ethical or practical to expect people with a long-duration infection to isolate.

Contact tracing has to a limited extent also been used to control influenza outbreaks [11, 12]. Contact tracing was a “critical intervention” in the Liberian Ebola epidemic of 2014-2015 and “represented one of the largest contact tracing efforts during an epidemic in history” [13].

This raises the question: How effective is contact tracing coupled with isolation (CTI) at controlling outbreaks of short duration illnesses such as those associated with coronaviruses and influenza? And how well implemented must contact tracing and isolation be? After all, CTI is potentially a far less costly and intrusive way of controlling dangerous illnesses than lockdowns, though both together may be necessary in some situations.

It’s impossible to provide a precise answer to this question. Infection outbreaks have practically infinite variations and are extraordinarily complex and stochastic. As the SARS-CoV-2 pandemic has shown, human populations are highly heterogeneous and it’s hard to predict how even a suburb, let alone a city or country, will be affected by an outbreak.

Nevertheless models, in particular agent-based ones, can provide insight into infectious disease dynamics [15, 16], and particularly the effectiveness of CTI versus isolation only of infected people versus no contact tracing or isolation at all. If simulations of human populations can show us that under a wide variety of assumptions, CTI is likely to be beneficial then it is worth implementing CTI in response to dangerous infection outbreaks.

## 2 Methodology

We simulated thousands of short duration illnesses with the following model world:

- A population of 10, 000 agents is initiated with ten randomly infected agents. The remaining agents are in a *Susceptible* state.
- Infected agents are initially in an *Exposed* state. They then advance to an *Infectious Asymptomatic* state. Then they advance either to an *Infectious Symptomatic* or *Recovered* state. An agent that has advanced to the *Infectious Symptomatic* stage advances to the *Recovered* state. ^1^
- Uninfected agents may become exposed if they are adjacent to one of *k* neighbouring agents in one of the two infectious states. Each uninfected agent has its own susceptibility to being infected by its neighbours. Each infected agents also has its own level of infectiousness.
- Agents are tested for the infection with a specified probability per day depending on the state they are in. The test result becomes known an average of a specified number of days later.
- Agents that test positive may be placed in isolation for a fixed number of days in which case they are less likely to become infected. Each agent has its own adherence level to isolation which is factored into its risk of being infected. There is a continuum of isolation adherence that affects the risk of infection.
- If an agent is one of the *k* adjacent neighbours of an agent whose test result is known it may be traced with a specified probability. If it is successfully traced it is also placed into isolation.
- We assume we are interested in highly contagious diseases with at most a single digit infection mortality rate, such as SARS-CoV-2 or influenza, and unlike MERS or Ebola (though our work can easily be extended to examine these as well). Hence we have not factored death into the simulations.

Thus the risk of infection of an uninfected agent, *a*, that comes into contact with an infectious one, *b*, is a stochastic function of the susceptibility to infection of *a*, the infectiousness of *b* and the isolation of *a* and *b*.

A simulation runs as follows: First the agents are initialized. Then for 500 iterations, where each iteration represents a day, the following events take place: new infections, testing, isolation, de-isolation (for agents who have been isolated for the specified number of days), tracing, and finally disease progression.

The simulation engine has the algorithmic form described in Listing 1.

Listing 1: Structure of the simulation engine

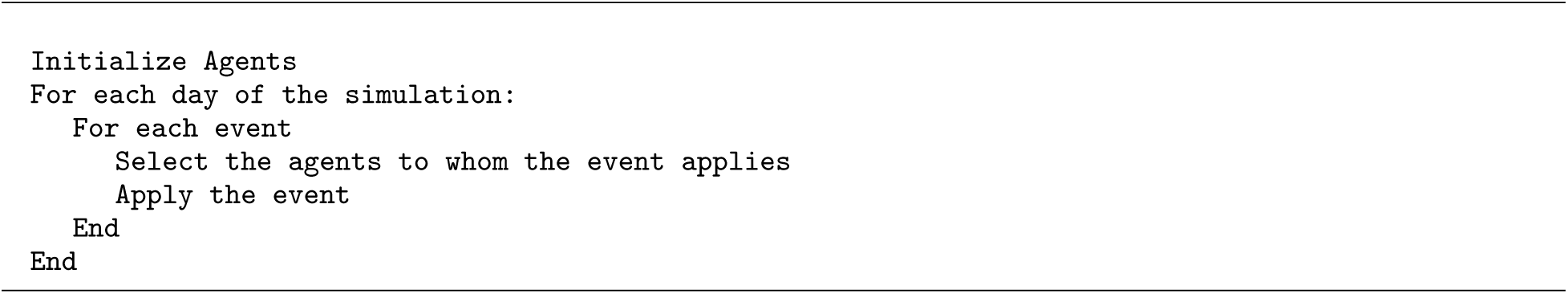

This structure is based on [14].

By design our model, named HETGEN, encompasses extremely heterogeneous and stochastic agent behaviour: (1) each agent has its own susceptibility when uninfected, and infectiousness when infected, (2) infected agents traverse through the infection stages stochastically, (3) agents have their own isolation adherence parameter, (4) agents get tested stochastically but infected symptomatic agents are much more likely to get tested, (5) tracing occurs when an infected agent’s test is returned with a positive result, and (6) tracing, too is stochastic, with a success probability for each contact.

Listing 2 is the pseudo-code for the infection algorithm.

Listing 2: Infection algorithm

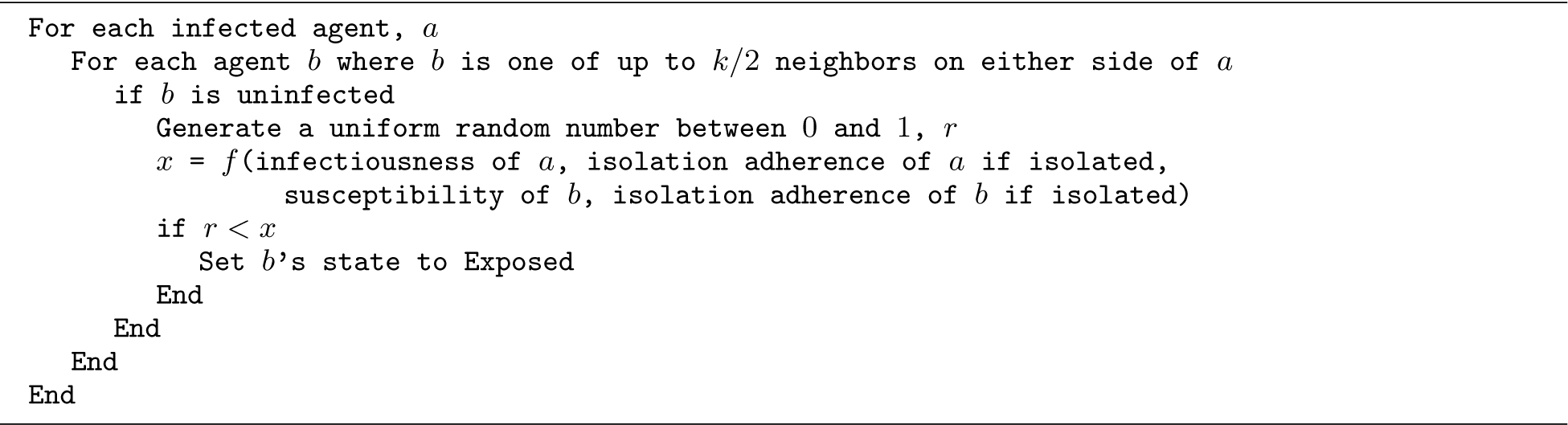

The infection function, *f* in Listing 2 for two agents, *a* and *b*, where *a* is infectious and *b* is susceptible, *i* is a property between 0 and 1 of *a* and *b* measuring their adherence to isolation, *t* is the infectious of *a*, and *s* is the susceptibility of *b*, is as follows:

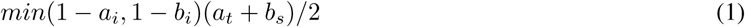

This unstructured model is designed to contrast, and complement, a highly structured model we have previously described that was designed to capture specific characteristics of Covid-19 and the effect of CTI on reducing SARS-CoV-2 infections in a South African township in which some agents attend schools, some work, some use taxis, and all live in households which are located in neighbourhood blocks, with each of these settings conferring different risks.[18]

### 2.1 Scenarios

We compared five scenarios:

**None** There is no isolation or contact tracing.

**Isolation** There is only isolation of agents with positive test results, with 85% mean adherence per day.

**Minimum** There is isolation of agents with positive tests results with 85% mean adherence per day, and 10% of their contacts are traced and isolated with 85% mean adherence per day.

**Moderate** There is isolation of agents with positive results with 85% adherence per day, and 30% of their contacts are traced and isolated with 85% mean adherence per day.

**Maximum** There is perfect isolation of agents when they test positive and perfect tracing of contacts of their contacts, who are then also perfectly isolated.

We ran 15, 000 sensitivity tests, repeated 10 times for each scenario, on 10, 000 agents. This comes to 15, 000 *×* 10 *×* 5 = 750, 000 simulations.

On each sensitivity test several of the parameters are randomly perturbed, using a uniform distribution, over a specified range. Table 1 lists the parameters used in the model. Entries in the value column separated by a hyphen are the perturbed parameters, while those that do no have a hyphen are held constant across all simulations.

**Table 1:** Parameters used in the simulations. Entries in the Value column with a hyphen are randomly perturbed before each sensitivity test.

| Parameter | Value | Notes |
| --- | --- | --- |
| Number of days | 500 | Each simulation iteration corresponds to 1 day |
| Initial infections | 10 | Infected agents are set to Exposed |
| Daily probability of Susceptible agent getting tested | 0 |  |
| Daily probability of Exposed agent getting tested | 0.1-0.3 |  |
| Daily probability of Infectious Asymptomatic agent getting tested | 0.1-0.3 |  |
| Daily probability of Infectious Symptomatic agent getting tested | 0.3-0.9 |  |
| Probability of exposed agent test returning positive | 0.5 |  |
| Probability of Infectious Asymptomatic agent test returning positive | 0.9 |  |
| Probability of Infectious Symptomatic agent test returning positive | 0.999 |  |
| Mean number of days for test result to come back | 1-6 |  |
| Minimum number of days for test result to come back | 0-2 | Even if mean is less than this, this prevails |
| Isolation days | 6-14 |  |
| Probability per day of staying Exposed | 0.1-0.9 |  |
| Probability per day of staying Infectious Asymptomatic | 0.1-0.9 |  |
| Probability of changing from Infectious Asymptomatic to Recovered | 0.05-0.95 |  |
| Probability per day of staying Infectious Symptomatic | 0.1-0.9 |  |
| Number of neighbours affected in matching and isolation | 32-44 |  |

### 2.2 Limitations

While the model encompasses a wide variety of variables for a wide number of possible illnesses, it is merely a computer simulation and cannot capture all real-world dynamics. Also the search space of possible illnesses, even with the limited number of variables we have perturbed, is massive (indistinguishable from infinity for practical purposes). 15, 000 illnesses represents a tiny fraction of the search space. Nevertheless our sample should be large enough to draw tentative conclusions that inform policy, or at least further clinical research.

A further limitation of the infection algorithm is that only the agents in the neighbourhood of an infectious agent can become infected. This algorithm, while more realistic than random or unassortative mixing ones, certainly doesn’t capture the complexity of real-world contacts. In future work we will explore compromises between assortative and unassortative mixing.

Also there is no migration by agents into or out of our model-world.

While this work may suggest that CTI can mitigate the spread of infectious disease, it offers no insight into how it can be implemented effectively.

### 2.3 Programming

Our model was prototyped in Python and then recoded, and further developed, in C++. This allowed us to run hundreds of thousands of simulations in several hours on affordable, standard consumer hardware. We recommend this methodology to other modellers who are welcome to use our code as a basis for this kind of simulation. The HETGEN source is available under the GNU General Public License version 3.0. Our code and results are available at https://github.com/nathangeffen/ABM_CTI.

### 2.4 Calibration, *R*_0_ and wide range of infections

To the extent that the model has been calibrated, it has been done experimentally. The values in Table 1 that are perturbed in 15, 000 sensitivity tests, have been set to go slightly beyond the range of reasonable estimates for influenza and SARS-CoV-2 infections. Those that are not perturbed have been set to what we hope are reasonable values.

Because a practically infinite number of illnesses are encompassed in these values, many outbreaks do not have an *R*_0_ above 1 and fizzle out immediately. Others, on the other hand, have extremely high *R*_0_ values, with every agent becoming infected if no intervention takes place. Calculating *R*_0_ per simulation for such models in which the agents behave so heterogeneously is also not useful, with different methodologies giving widely different estimates. We therefore do an analysis of the results that includes all the simulations and sub-analyses that exclude those where fewer than 500 agents become infected.

## 3 Results

We calculated the mean, median and standard deviation over the ten runs of each of the 15, 000 sensitivity test for each of the five scenarios. Table 2 shows the results.

**Table 2:** Results of the 15, 000 sensitivity tests using 10, 000 agents.

| Scenario | Mean | Median | Std |
| --- | --- | --- | --- |
| None | 5,743 | 7,511 | 4,269 |
| Isolation | 5,341 | 5,983 | 4,285 |
| Minimum | 4,419 | 2,991 | 4,092 |
| Moderate | 3,359 | 1,282 | 3,731 |
| Maximum | 994 | 273 | 1,716 |

As expected, the *None* scenario had the highest mean and median total infections, followed by *Isolation, Minimum, Moderate* and *Maximum*.

Note that the median for the *None* scenario is higher than the mean, indicating that the mean is pulled down disproportionately by illnesses with low *R*_0_ that never become sizeable. By contrast the medians of the three most effective intervention scenarios are lower than the means, indicating that the means are increased disproportionately by epidemics with large *R*_0_ that, despite the intervention, still resulted in a large number of infections.

Also useful is to know for how many illnesses each scenario outperformed the others. We compared scenarios using the mean of the ten runs for each illness. If an intervention has no effect, it should be outperformed by the None scenario approximately 7, 500 times (i.e. half of the 15, 000 illnesses). Using the mean of the ten runs per illness, the None scenario only outperformed the Isolation, Minimum, Moderate and Maximum interventions 2, 986, 828, 423 and 193 times respectively. (Note: Whenever the None scenario beats another scenario, it is only due to the stochastic nature of the simulations.)

Many illnesses never reach epidemic proportions or the difference between the interventions is small. We therefore did a further analysis of counting the number of times each scenario outperformed (or was outperformed by) the no intervention scenario if two criteria were met: (1) A mean of at least 500 agents were infected in the None scenario across the tens runs of an illness and (2) the better performing scenario had no more than 70% of the infections of the scenario it was being compared to. Using these criteria the None scenario only outperformed the Isolation, Minimum, Moderate and Maximum interventions 28, 2, 0 and 0 times respectively. It was outperformed by them 2, 020, 5, 406, 7, 754 and 11, 392 times respectively. Making the second criterion even more stringent, by lowering it to 40%, resulted in the None scenario never outperforming any of the intervention scenarios. The intervention scenarios outperformed the None scenario 346, 2, 599, 5, 657 and 10, 585 times respectively.

## 4 Conclusions

Our analysis suggests that if an infectious outbreak occurs with certain characteristics similar to influenza and SARS-CoV-2, even moderately well implemented CTI is likely to substantially reduce the ultimate number of infections. These characteristics are: (1) There must be a test for the illness with reasonable specificity and sensitivity for which results can be obtained in a few days. (2) A significant number of infectious people should present for the test, (3) the length of the illness should exceed the mean turnaround time for the test, and (4) the means to carry out contact tracing and isolation must exist. These characteristics are achievable for seasonal influenza and SARS-CoV-2 in many countries.

Even moderately implemented isolation measures without tracing are likely to have a substantial benefit. This suggests that if people are, for example, strongly encouraged to stay at home if they have influenza symptoms during the influenza season, many infections, and consequently a substantial number of deaths, can be averted, even more so perhaps if entire households remain at home while one member is symptomatic with influenza. It’s conceivable that this may even reduce the total number of days absent from work during influenza season, though further research is needed to test this hypothesis.

Randomized cluster-controlled clinical trials conceivably could test the efficacy of contact tracing. A Cochrane Review has pointed out the lack of contact tracing randomised trials for tuberculosis [17]. This gap in medical evidence applies to most infectious diseases. While models such as ours suggest that it is worthwhile investing in contact tracing infrastructure for infectious diseases, randomised trials can offer much clearer insight into the cost-effectiveness of CTI and its practical feasibility than models.

Interestingly, because our model is highly heterogeneous and our analysis finds that, occasionally, even with the best possible implementation of CTI, there will be no benefit merely due to stochastic effects. We suspect this is a finding with real-world relevance. It is likely that in two similar settings, wherein no intervention is implemented in one, while a comprehensive CTI intervention is implemented in the other, the latter will, in a minority of occasions, have a worse epidemic, solely due to the stochastic nature of epidemics.

## Data Availability

All code and data are available on Github.

https://github.com/nathangeffen/ABM_CTI

## Footnotes

1 While our simulation program can handle hospitalisation, intensive care and death states, we decided that this was unnecessary complexity for our purposes here.

